# PRAYAS: Cohort profile for Pooled Research and Analysis for Yielding Anemia-free Solutions in India

**DOI:** 10.1101/2025.08.07.25333266

**Authors:** Anuj Kumar Pandey, Anju Pradhan Sinha, Ramu Rawat, Ranadip Chowdhury, Shivaprasad S Goudar, Jitender Nagpal, Shrey Desai, Avula Laxmaiah, Kalpana Basany, Sadhana Joshi, Chittaranjan Yajnik, Aparna Mukherjee, Pratibha Dwarkanath, Priyanka Gupta Bansal, Molly Jacob, Shinjini Bhatnagar, Komal Shah, Debarati Mukherjee, Amlin Shukla, Raghu Pullakhandam, Varsha Dhurde, Aditi Apte, Rajeev Singh, Aakriti Gupta, Pearlin Amaan Khan, Usha Dhingra, Ravi Upadhyay, Sutapa Bandyopadhyay Neogi, Manjunath S Somannavar, Anirban Mandal, Gayatri Desai, S Sengupta, Shailendra Dandge, Girija Wagh, Urmila Deshmukh, Gunjan Kumar, Anura V Kurpad, G.S. Toteja, Nikhitha Mariya John, Shailaja Sopory, Somen Saha, Giridhar R Babu, Anandika Suryavanshi, Ravinanadh Palika, Archana Patel, Radhika Nimkar, Gaurav Raj Dwivedi, Umesh Kapil, Yamini Priyanka, Arup Dutta, Sunita Taneja, Diksha Gautam, Avinash Kavi, Swapnil Rawat, Kapilkumar Dave, R Raman, Catherine L. Haggerty, Sajay Lalwani, Prachi Phadke, Alka Turuk, Tinku Thomas, Neena Bhatia, Manisha Madai Beck, Lovejeet Kaur, Aakansha Shukla, R Deepa, Lindsey M Locks, Dhiraj Agarwal, Raja Sriswan Mamidi, Harshpal Singh Sachdev, Rounik Talukdar, Sayan Das, Nita Bhandari, Ranjana Singh, S YogeshKumar, Ramasheesh Yadav, PS Reddy, Sanjay Gupte, S. Rasika Ladkat, Zaozianlungliu Gonmei, Swati Rathore, Dharmendra Sharma, Apurvakumar Pandya, Yamuna Ana, Patricia Hibberd, Himangi Lubree, Anwar Basha Dudekula, Priti Rishi Lal, Dilip Raja, Aruna Verma, Umesh S Charantimath, Meshram, Karuna Randhir, Onkar Deshmukh, Ashok Kumar Roy, Obed John, Nolita Dolcy Saldanha, Ashish Bavdekar, Raj Kumar, Shyam Prakash, Wafaie W. Fawzi, Sunil Sazawal

## Abstract

**Purpose:** This cohort would aim to estimate the prevalence of anemia among children under 18 years, non-pregnant and non-lactating (NPNL) women, and pregnant women (by trimester), with further stratification by age group, year, and region of India. Cohort would also help address extended deliberation concerning etiological fraction of iron and other key erythropoietic micronutrient deficiencies contributing to anemia in India. Additionally, this will help assess the effectiveness of existing anaemia prevention and treatment interventions and examine factors associated with non-response, thereby supporting the “test–treat–track” approach.

**Participants:** Children under 18 years, pregnant women, and non-pregnant-non-lactating women (NPNL) in India.

**Findings to date:** This cohort profile comprises 88 datasets spanning between 1994 to 2023, encompassing a total of 319,721 participants for prevalence analysis [children(19,762), NPNL(17,883), and pregnant women(282,076)]. Additionally, 59,292 participants were included in intervention studies [children(13,435), NPNL(11,594), and pregnant women(34,263)]. RCTs comprised 55.7% (49/88) of the datasets whereas observational studies comprise 35.2% (31/88) of the datasets. Majority of the studies were from the norther region - 38 studies (43.2%), followed by the western part - 20 studies (22.7%). The southern part contributed 16 studies (18.2%). Major [59/88 (67%)] datasets in cohort were from community-based studies. The sample included NPNL and pregnant women with a median age of 26 years (IQR 23-32), and 23 years (IQR 21-25) respectively. Information from 6 months up to 18 years was pooled within the children’s cohort. Within the pregnancy cohort the mean gestational age at enrollment was 10.24 weeks(SD-17.65). Of total 10.8% (34,442/ 319,721), 9% (28,672), 4.5% (14,240) of the sample had information on complete blood count, ferritin and vitamin B12 respectively. A total of 33 datasets (sample - 59,292) were from intervention studies. Among pregnant women, a broader range of interventions was implemented, including intravenous iron sucrose, ferric carboxymaltose, iron isomaltoside, IV iron combined with vitamin B12, folic acid, and niacinamide, integrated interventions, as well as low-dose calcium supplementation. A similar set of interventions were delivered to NPNL group with being distinct which compared Ferrous sulfate tablets of 60 mg elemental iron daily with a control of 120 mg on alternate days. Ferrous sulfate was the major interventions amongst children along with food supplements and some were Ayush trials.

**Future plans:** The PRAYAS will provide robust, high-quality evidence to inform public health policy in India. The findings will feed into the Anemia Mukt Bharat program recommendations for pregnant women, NPNL women and children to guide targeted strategies for reduction of anemia and its associated health burdens across vulnerable populations.

**Strengths and limitations of this study:**

1. The harmonized PRAYAS pooled Indian dataset is one of the largest, reliable and most comprehensive datasets on pregnant/ non-pregnant and non-lactating women and children.
2. One of its kind of dataset with information on hemoglobin levels, relevant biochemical and key micronutrients parameters and varied interventions from across India.
3. Heterogeneity of interventions, dosage, duration and data collection approaches.
4. Studies lack critical parameters needed to assess changes in haemoglobin concentration like non-availability of key erythropoietic micronutrients in most of the studies, limiting the scope of certain analyses.

## Introduction

Nutritional anemia remains a significant global public health challenge (1,2), with profound implications for the health and productivity of women and children. In 2019, anemia affected nearly one-third of women of reproductive age (WRA) globally, with approximately 269 million children aged 6–59 months also impacted (3). The burden of anemia is disproportionately high in low- and middle-income countries (LMICs) (4), with the African and South-East Asian regions contributing the most to global prevalence (5,6). In LMIC, anemia affects 43% of the population compared to just 9% in developed nations, with WRA and children being the most vulnerable groups(7,8). While anemia has a multifactorial etiology, iron deficiency is the most prevalent cause, particularly in LMICs, where nutritional deficiencies are widespread (9). The World Health Organization (WHO) Global Nutrition Targets aim for a 50% reduction in anemia among WRA by 2030, reflecting its prioritization in global health initiatives (10).

In India, the burden of anemia has shown alarming trends. The National Family Health Survey (NFHS-5) highlights an increase in anemia prevalence among WRA (from 53.1% in 2015–16 to 59.1% in 2019–21), pregnant women (from 50.3% to 52.6%), and children aged 6–59 months (from 58.6% to 67.1%) over the same period (11). However, the methods used for assessments and the interpretation of the results have highlighted several challenges, like the use of capillary blood, unlike gold standard methods of assessments through venous blood (12–14). Additionally, a national survey reported that ∼41% of preschoolers, school-age children, and adolescents (aged 1–19 years) were anemic, with female adolescents experiencing higher prevalence rates (40%) than males (18%) (15). Anemia’s consequences are far-reaching, including fatigue, impaired cognitive and immune function, reduced productivity, and increased morbidity and mortality(16,17). Addressing anemia remains a public health priority in India, as evidenced by initiatives like the Anemia Mukt Bharat (AMB) program, launched in 2018 to reduce anemia prevalence by three percentage points annually among children, adolescents, and WRA(17). The reduction of anemia is one of the important objectives of the POSHAN Abhiyaan launched in March 2018.

Despite these efforts, the prevalence of anemia remains unacceptably high (18,19). Improving the understanding of anemia’s burden across demographic groups and evaluating the effectiveness of interventions are critical for guiding policy and program decisions. Complying with the targets of POSHAN Abhiyaan and National Nutrition Strategy set by the NITI Aayog, AMB strategy has been designed to reduce prevalence of anemia.

There is a need to synthesize the evidence on anemia and analyze the progress made under AMB or reasons for its inadequate progress. In this context it was decided to collate all existing data on anemia from studies conducted over more than four decades across diverse regions of the country by contacting the investigators.

Primary goal of this initiative is to answer the questions that remain unanswered by individual studies. The results would feed into the AMB program recommendations for WRA and children. The results would also guide targeted strategies to reduce anemia in India. It is expected that by pooling the observational and interventional studies we may investigate the etiological fractions of various causes of this recalcitrant public health problem and synthesize evidence on effectiveness of several interventions used for prevention and treatment of anemia. By integrating data from a wide range of geographical, community, and healthcare settings, this extensive pooling of studies aims to provide a comprehensive and nuanced understanding of the underlying trends and patterns. Our approach not only enhances the depth and breadth of available evidence but also ensures a more representative and holistic perspective on the factors influencing health outcomes over time.

### Cohort Description

#### a) Selection of studies

Given the persistent and high burden of anemia in India despite ongoing initiatives, there was a recognized need for an in-depth understanding of the issues concerned with anemia across the country. In response, the <u>Director General of the Indian Council for Medical Research (ICMR) commissioned an initiative in early 2023</u> to conduct a comprehensive assessment of the anemia burden in India. To facilitate this effort, ICMR constituted a committee of eminent experts in the field of anemia in India. The initial phase involved key preparatory activities, including the development of database search keywords, identification of principal investigators (PIs) and relevant studies across India, as well as contacting the study PIs. These activities were undertaken by the Secretariat at the ICMR.

A designated committee conducted the selection of studies under the chairperson’s guidance. To identify relevant studies, committee adopted two well-established approaches. First, a systematic search of trial registries, and second, collaborative discussions with investigators of ongoing studies involving children and women (pregnant and NPNL) (20,21). The process began with a database search of the Clinical Trials Registry of India (CTRI)(22). The initial search was conducted using designated keywords such as ‘anemia’, ‘prevalence’, ‘children’, ‘randomized controlled trial’, ‘intervention studies’, ‘anemia etiology’, ‘pregnant women’, ‘women of reproductive age’, and ‘non-pregnant women’(22).

Committee also expanded the search by examining cross-references from related studies, following up with leads provided by principal investigators (PIs) of studies included, and reaching out to additional researchers in the field. After identifying related studies, committee members contacted the PIs with requests for collaboration in early-2024. Once the PIs agreed, a data sharing agreement was prepared and signed by them, and several online meetings were held to discuss the details of the proposed cohort and the datasets involved. After the online meetings with study PIs, the harmonization meeting was held at ICMR headquarters in New

Delhi in early August 2024 with the objective of discussing methodological issues, explaining to the participants what the cohort would be and discussing the difficulties in filling the data extraction sheets. Such meetings were scheduled at the ICMR headquarters at routine intervals of 3-4 months’ time. The meeting also aimed at handholding the PIs on how to fill the data cells. A second round of data harmonization meeting was held at the end of October 2024. After identifying and removing duplicates, <u>a total of 88 datasets from studies,</u> <u>conducted by 23 organizations at different time points in India were included for the cohort synthesis</u> <u>following certain inclusion criteria</u> -

- This cohort included studies on anemia conducted on children under 18 years, NPNL women, and pregnant women, where data on hemoglobin levels were available.
- Eligible studies included cross-sectional and interventional designs (both randomized and non-randomized), longitudinal studies, unpublished studies, including the COVID registries with due approval from the relevant authorities.
- Studies conducted in India, with data on hemoglobin and other relevant biochemical parameters (e.g., serum ferritin levels, complete blood counts, vitamin B12 levels, inflammatory markers) at baseline and post intervention were also included within the cohort.

Data sets were excluded if data on hemoglobin and other relevant biochemical parameters were not available.

#### b) Data sources in the pooled data set

After completing the formalities and harmonization, the PIs shared their anonymized datasets in a predefined format. The data included information such as study ID, woman/subject ID, demographic details, interventional strategies, hemoglobin and ferritin levels, relevant biochemical parameters at baseline and post-intervention, comorbidities, adverse effects, etc.). Figure 1 provides details of the studies identified through databases and number of studies included in the final cohort. The details of each of the studies included are available elsewhere (Supplementary File-S-Table 1).

**Figure I:**
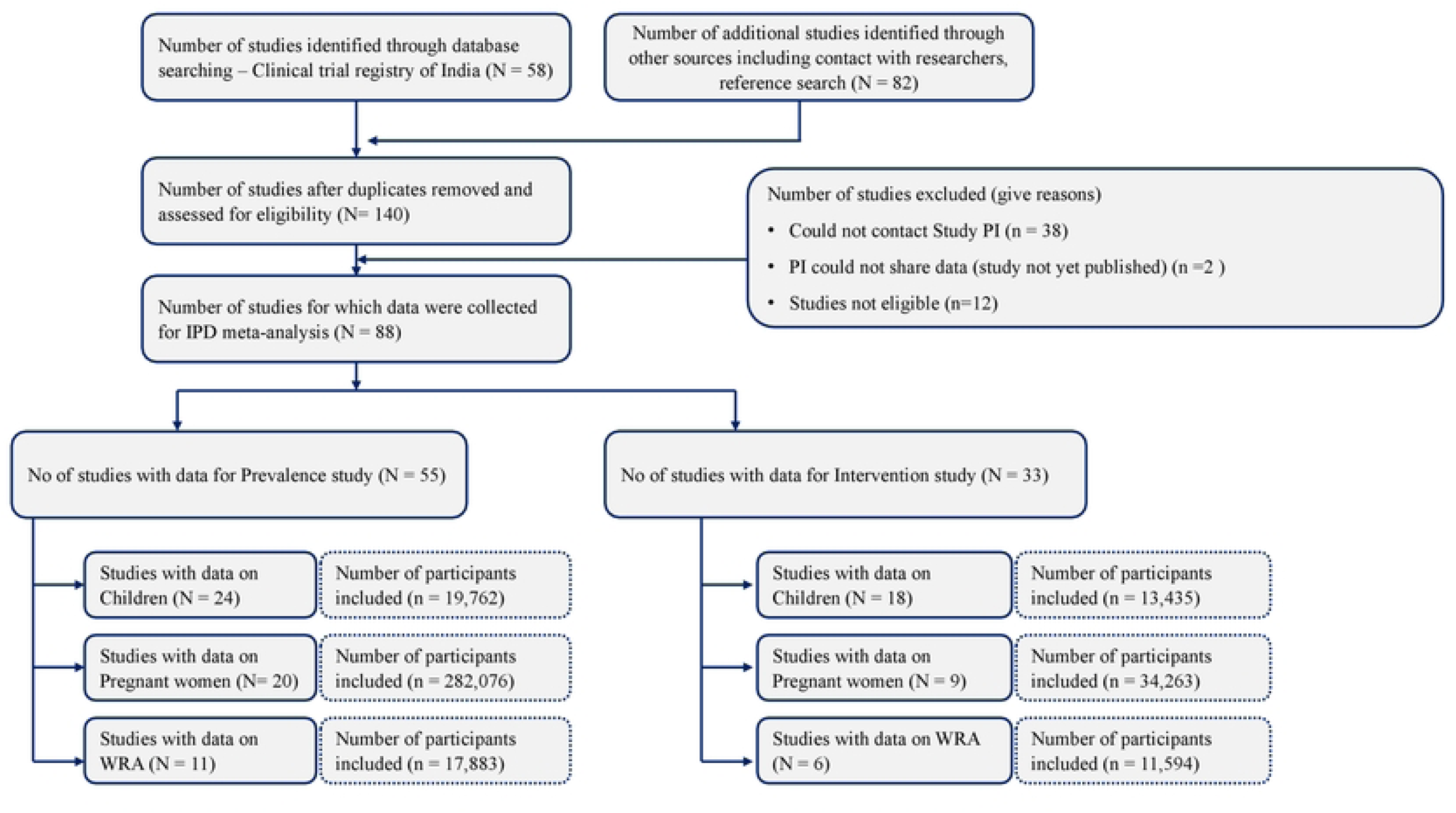
Overvie,v of the data included in the Pooled Cohort.

The ICMR team ensured that the included studies complied with relevant ethical guidelines and regulations. All included primary studies had received approval from ethics committees recognized by the Department of Health Research in India, and informed consent was obtained from all study participants. Table 1 provides details of studies included within the cohort profile of PRAYAS.

After finalizing the datasets, the entire cohort was separated into groups for <u>children under 18 years, NPNL, and</u> <u>pregnant women</u>. The studies were then categorized into two groups:

1. Prevalence studies and
2. Intervention studies

**Table 1:**
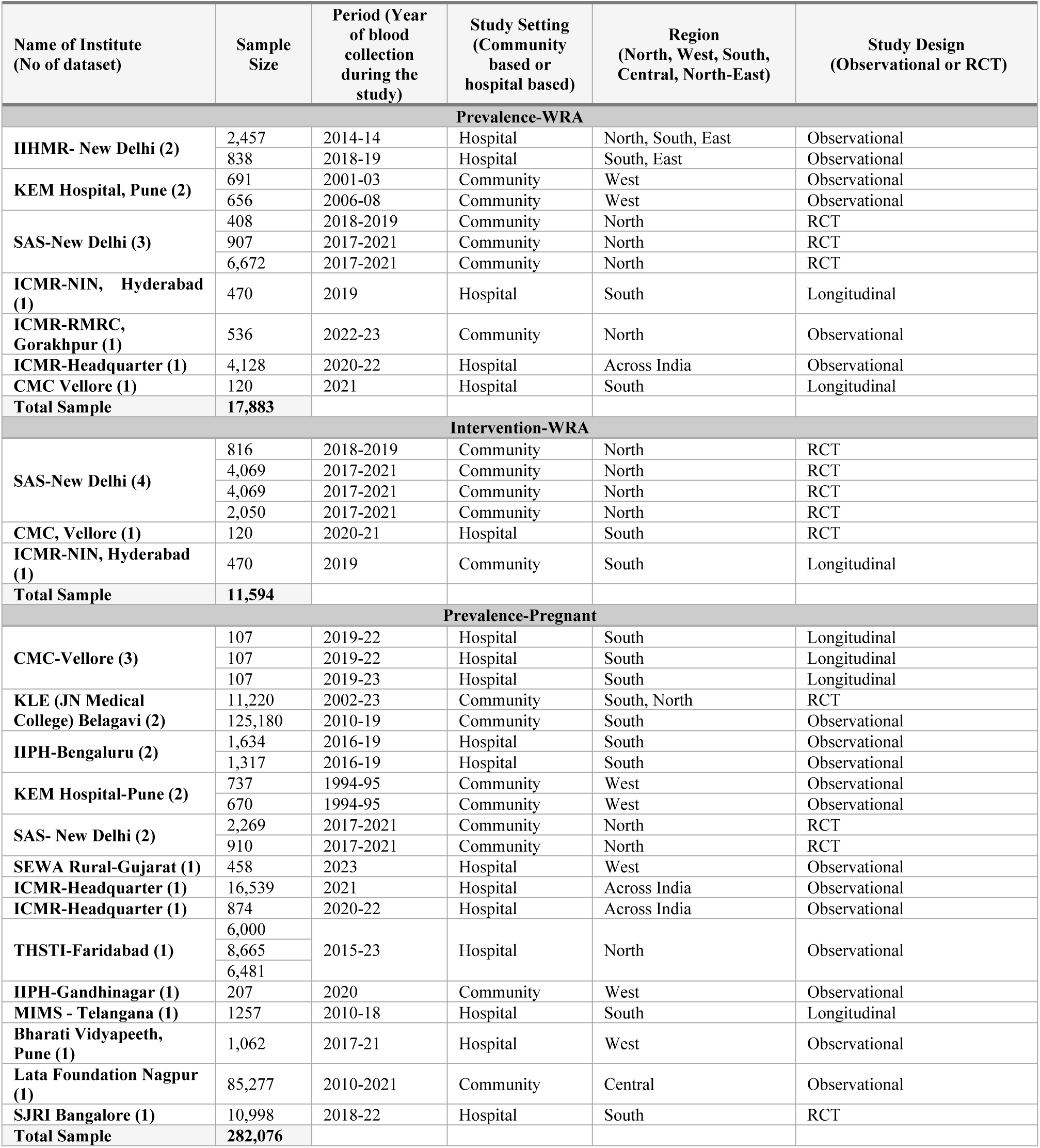
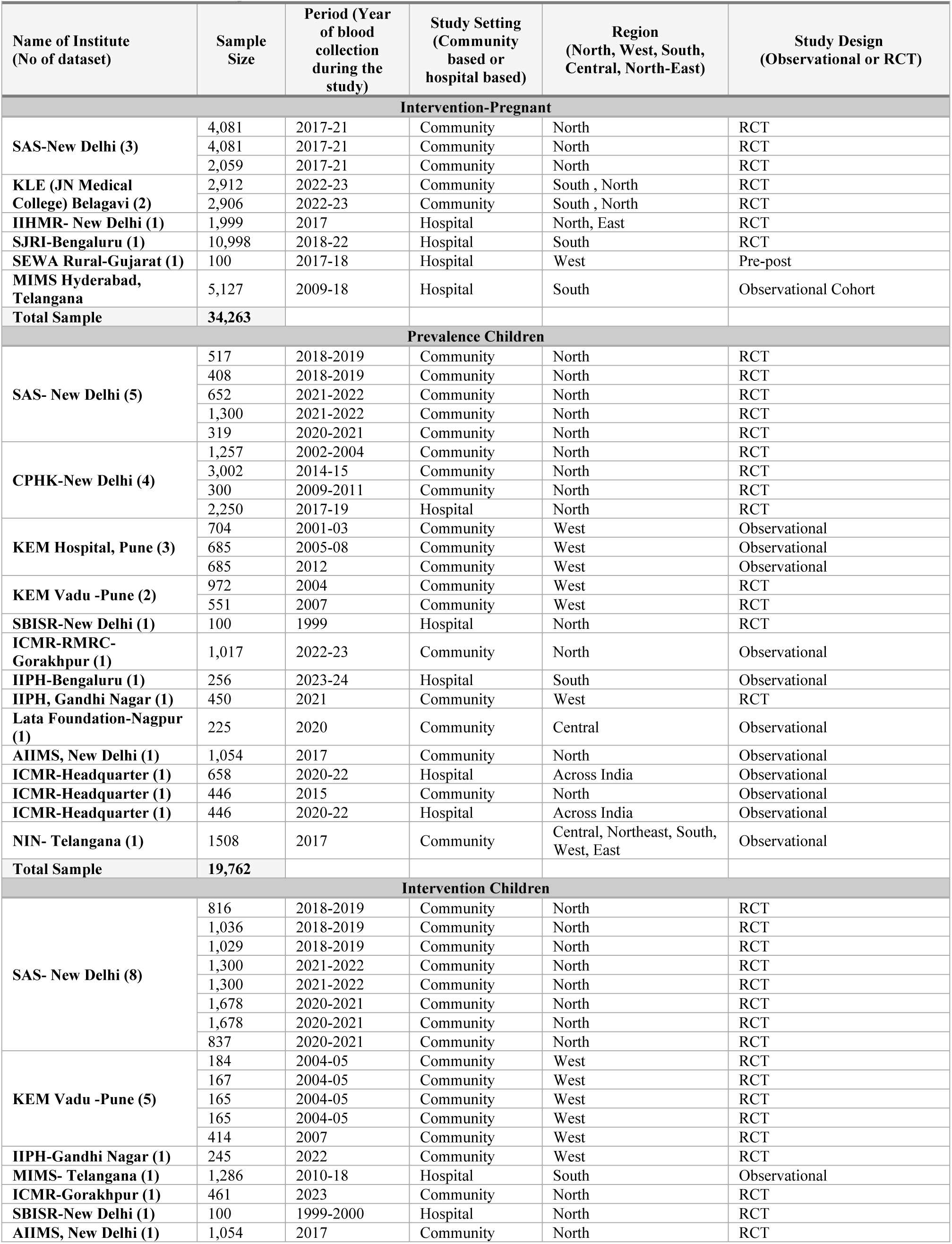
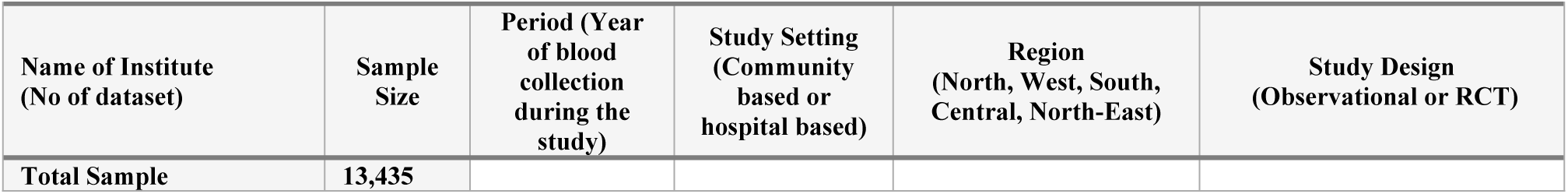
Details of studies – cohort profile PRAYAS.

This categorization was important since it dictated the type of analysis and statistical methods to be applied. The included studies (Table 1) span both hospital and community settings and include observational, longitudinal, randomized controlled trials (RCTs), and pre-post intervention designs. States and UTs were classified as regions for analysis following the classification system set by the Registrar General & Census Commissioner of India for sample registration system (SRS) (23).

#### c) Variable availability and definition

Data harmonization is a critical step while developing a cohort profile, ensuring that data from diverse studies can be integrated and analyzed collectively, thus enhancing the reliability and generalizability of the findings (24–26). A significant aspect of harmonization was to ensure uniform units for all biochemical variables. For example, hemoglobin levels (reported in grams per deciliter or grams per liter by different studies), were standardized to a single unit (grams per deciliter). This step, along with the standardization of other blood parameters such as red blood cell count, and serum ferritin etc., was also undertaken. A separate sheet with standardized parameters was developed for reference (Table 2).

**Table 2:**
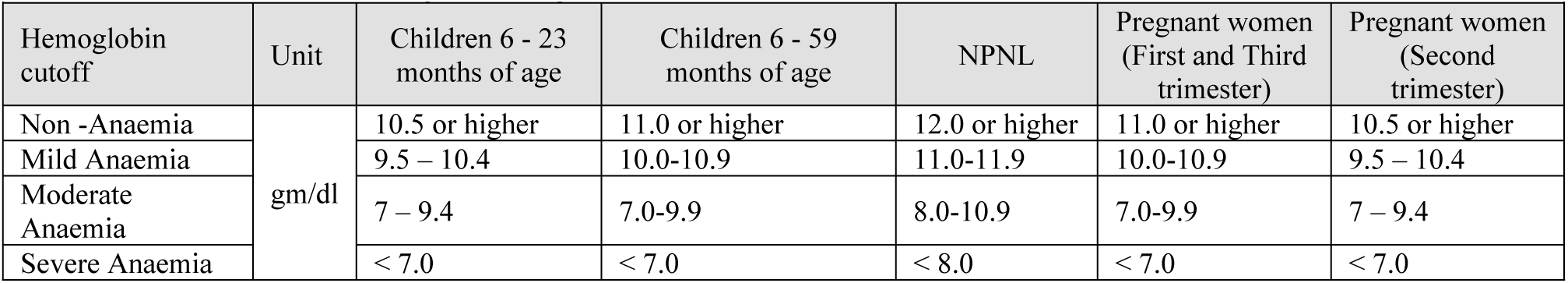
Cut off values for haemoglobin along with the unit for data collection.

Hemoglobin (Hb), the primary outcome indicator of anemia, was measured in grams per deciliter (g/dL), and categorized as mild, moderate and severe based on the hemoglobin threshold as mentioned in the updated guideline on haemoglobin cutoffs to define anaemia, released in 2024 (Table 2) (27,28).

Table 3 also presents the acceptable upper and lower values for each hematological and biochemical biomarkers for children, pregnant and non-pregnant women. These values served as quality control measures to exclude implausible values. Additionally, the table also presents the acceptable unit for each parameter. Definition for the micronutrient-related thresholds, inflammatory and metabolic markers were also defined to check for the quality of collected data.

**Table 3:** Acceptable values to eliminate abnormal values from the cohort.

|  |  |  |  |  |  |  |
| --- | --- | --- | --- | --- | --- | --- |
| Parameter | Children |  | Pregnant women |  | Non- pregnant women |  |
|  | Lower acceptable value | Upper acceptable value | Lower acceptable value | Upper acceptable value | Lower acceptable value | Upper acceptable value |
| Hematocrit (29) | <30% | >44.1% | <36% | >48% | <36% | >48% |
| MCV(30) |  | > 86 femtoliter (fL) | <80 femtoliter (fL) | > 100 femtoliter (fL) | <80 femtoliter (fL) | >100 femtoliter (fL) |

|  |  |  |  |  |  |  |
| --- | --- | --- | --- | --- | --- | --- |
| **Parameter** | **Children** |  | **Pregnant women** |  | **Non- pregnant women** |  |
|  | Lower acceptable value | Upper acceptable value | Lower acceptable value | Upper acceptable value | Lower acceptable value | Upper acceptable value |
| MCH (31,32) | (6m-1 yr)23 pg | (6 m-1 yr) 31 pg | > 33 picograms (pg) per cell |  | > 33 picograms (pg) per cell |  |
| MCHC (33) | (6m-1 yr) <32 g/dL | (6m-1 yr) >36 g/dL | <32 g/dL | >36 g/dL | <32 g/dL | >36 g/dL |
| Ferritin (34) (35) |  | 140 µg/L | 13 µg/L | 150 µg/L | 13 µg/L | 150 µg/L |
| Transferrin Saturation (36–38). | (0 - <1 year) 4.1% | 30% (0 - <1 year) 59 % | 15% | 50% | 15% | 50% |
| sTfR (39). |  |  |  | 4.4 mg/L |  | 4.4 mg/L |
| ZnPP (39,40). |  | (Abnormal values) 3-6 years >70 µmol/mol heme |  | 100 µg/dL |  | 100 µg/dL |
| Vitamin A (42,43) | <0.70 µmol/L |  | 0.07 µmol/g | 3000 retinol activity equivalents (RAE)/Day | 0.07 µmol/L | 3000 retinol activity equivalents (RAE)/Day |
| Vitamin B12 (44,45) | <150 pmol/L (203 pg/mL) |  | 100 pmol/L | 350 pmol/L | 100 pmol/L | 350 pmol/L |
| Folate (Serum) (46,47) | < 4 ng/mL (<10 nmol/L) |  | 2.0 ng/mL | 7.0 ng/mL | 2.0 ng/mL | 7.0 ng/mL |
| Folate (RBC) (47,48) | < 151 ng/mL (<340 nmol/L) |  |  | >400 ng/mL |  | >400 ng/mL |
| Zinc (49) | <10 years:65 mg/dL |  | <10 years:65 mg/dL | 70 mcg/dL | <56 (µg/dL) | 70 mcg/dL |
| Vitamin D (50–52) | <12ng/mL |  | <30 nmol/L |  | 10 ng/mL | 50 ng/mL |
| CRP (53) | > 5 mg/L |  | 0.1 mg/L | > 5.0 mg/L | 0.1 mg/L | > 5.0 mg/L |
| AGP (54) | > 1 g/L |  | 0.4 mg/mL | 3 mg/mL | 0.4 mg/mL | 3 mg/mL |
| IL-6 (55) |  |  | 5 pg/mL | 25 pg/mL | 5 pg/mL | 25 pg/mL |
| D-Dimer (56) |  |  | 500 ng/mL | 10,000 ng/mL | 500 ng/mL | 10,000 ng/mL |

Note: RDW – Red Cell Distribution Width ; MCV – Mean Corpuscular Volume ; MCH – Mean Corpuscular Hemoglobin ; MCHC – Mean Corpuscular Hemoglobin Concentration ;sTfR – Soluble Transferrin Receptor; ZnPP – Zinc Protoporphyrin ;CRP – C-Reactive Protein ;AGP – Alpha-1-acid Glycoprotein ; IL-6 – Interleukin-6

#### d) Principles and plans for statistical analysis

A detailed statistical analysis and reporting plan was formulated in collaboration with the Technical Advisory Group of the PRAYAS consortium. This plan delineated the statistical techniques, underlying assumptions, and procedural steps, ensuring systematic and transparent analyses (details will be reported in subsequent papers). One-stage meta-analysis and two-stage meta-analysis approach would be used for analysing all the available data to calculate the prevalence of anemia and its severity across age groups. Using a weighted sample, the prevalence of anemia and its severity will be calculated as the number of anaemics divided by the total number of participants in different age groups. To account for differences in sample sizes across cohorts, each cohort will be weighted, with *weights computed as the inverse of the ratio of the individual cohort sample size to the overall pooled cohort sample size*.

The analysis will use a one-stage individual participant data meta-analysis approach, pooling harmonized data from all included studies to estimate the adjusted etiological fractions of anaemia due to specific micronutrient deficiencies (such as iron, folate, vitamin B12, vitamin A, vitamin D and zinc) in children, non-pregnant/non-lactating women, and pregnant women (by trimester where possible). Multilevel regression models will be employed, with study as a random effect and relevant covariates included to account for confounding and between-study heterogeneity. Adjusted risk ratios for each deficiency will be used to calculate PAFs, with subgroup analyses by age, region, and other modifiers. Sensitivity analyses will assess the robustness of findings to different deficiency cut-offs and model specifications. To evaluate intervention effects, logistic regression will estimate relative risks (RR) for anemia prevalence, while linear regression will compute mean differences (MD) in hemoglobin levels, adjusted for confounders. Additionally, the cohort would also be utilized to develop risk prediction models using machine learning approaches.

#### e) Patient and public involvement

No patients or members of the public were directly involved in the design or conduct.

### Findings to Date

The PRAYAS cohort, spanning over 379,013 individuals, offers a rich, regionally diverse, and methodologically varied resource to derive meaningful insights into nutritional anemia across India.

#### a) Study profile

Pooled cohort comprises 88 datasets, encompassing a total of 319,721 participants for prevalence analysis - children (19,762), NPNL (17,883), and pregnant women (282,076). Additionally, 59,292 participants were included in intervention studies - children (13,435), NPNL (11,594), and pregnant women (34,263). RCTs comprised 55.7% (49/88) of the datasets whereas observational studies comprise 35.2% (31/88) of the datasets. Others were longitudinal studies (8% - 7/88) and pre-post study (1.1% - 1/88). The included studies were conducted across various regions of India : the major contribution were from the norther region of India with 38 studies (43.2%), followed by the western part of India with 20 studies (22.7%). The southern part contributed 16 studies (18.2%). A smaller share from central part of the India (2.3% -2/88) followed by 12 studies (13.6%) from across India or have spanned multiple regions. These studies span from 1994 to 2023. A majority [59/88 (67%)] of the datasets originates from community-based studies, while 29/88 (33%) derived from hospital-based research (Table 1).

#### b) Baseline characteristics-Prevalence datasets

Of the included studies, more than 85% of the sample had information on hemoglobin concentration with highest amongst the pregnant women datasets with information from 96.1% (270,939) sample followed by NPNL and children with 94.4% (16,878) and 87.8% (17,351) respectively. The sample included NPNL and pregnant women with a median age of 26 years (IQR 23-32), and 23 years (IQR 21-25) respectively. Within the children datasets information from 6 months up to 18 years was pooled within the cohort. Ultrasonography was used in 76.8% (198,819) of the sample for gestational age assessment (23.2%-60,053 used LMP method). The mean gestational age at enrollment was 10.24 weeks (SD - 17.65). Specifically, more than one-third (41.32% - 105,103) of the participants were enrolled in the first trimester of pregnancy whereas 37% (94,249) in the second trimester.

Further assessment of information on each hematological and biochemical biomarker reported that overall, 10.8% (34,442/ 319,721) of the sample had information on Complete Blood Count (CBC). Whereas of the total sample, 9% (28,672), 4.5% (14,240) had information on ferritin and Vitamin B12 respectively. Less than 5% of the sample had information on other essential parameters (Figure 2). We are yet to analyze other essential parameters availability within the cohort.

**Figure 2:**
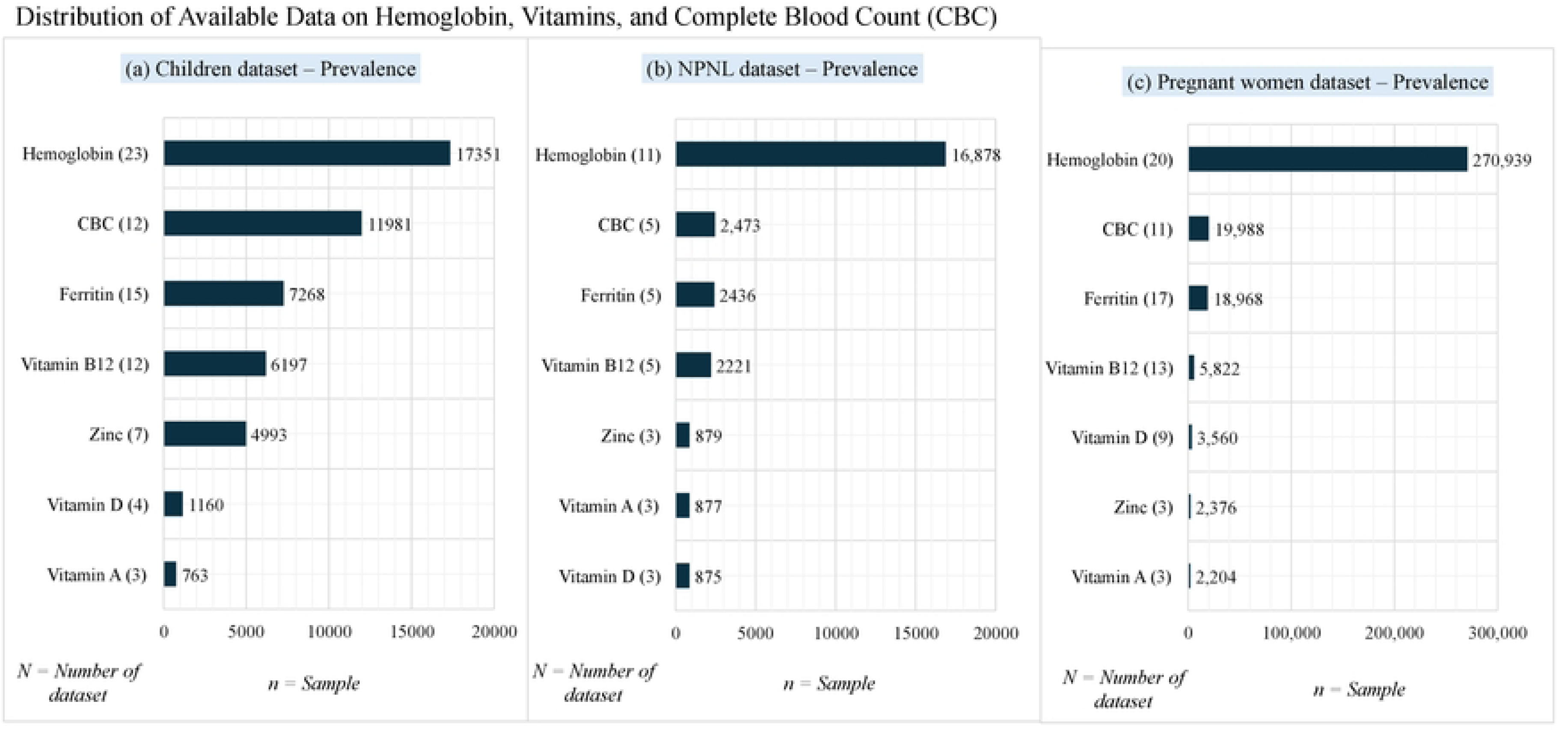
Distribution of Available Data on He1noglobin, Vitamins, and Co1nplete Blood Count (CBC) : (a) Children dataset ; ’NL dataset; (c) Pregnant women dataset

#### c) Baseline characteristics-intervention datasets

Within the PRAYAS cohort a total of 33 datasets (sample - 59,292) were from intervention studies. Of these, 87.9% (29/33) datasets are randomised controlled trials with maximum within the children cohort (17 datasets). This dataset focuses on addressing anaemia through nutritional and therapeutic approaches.

For the pregnancy cohort 23.8% (8,150/34,263) of the interventions were specified as therapeutic. It is pertinent to note that 54.6% (18,719) of the samples were not specified within the category of therapeutic or preventive. Among pregnant women, a broader range of interventions was implemented, including intravenous iron sucrose(18), ferric carboxymaltose (IV FCM) (57), iron isomaltoside (IV IIM) (57), IV iron combined with vitamin B12, folic acid, and niacinamide, integrated interventions (a combination of health, Nutrition, psychosocial care and WASH) (58,59), as well as low-dose calcium supplementation (60). These were administered either during pregnancy alone, during both preconception and pregnancy, or in the preconception period only (58,59). The control groups primarily received either high-dose calcium in one study(60) or oral iron in rest others.

For the WRA group 53.9% (6246 / 11,594) interventions were categorized as therapeutic. A total of 4 study namely WINGS (58,59), IMPRINT(61), ICMR NIN study(62), CMC-RCT contributed to the cohort. WINGS provided integrated interventions (a combination of health, Nutrition, psychosocial care and WASH). These interventions were delivered at different stages namely during preconception, during preconception + pregnancy and during pregnancy with a control of oral iron. Study by CMC compared Ferrous sulfate tablets of 60 mg elemental iron daily with a control of 120 mg on alternate days. Whereas NIN study administered prophylactic IFA and assessed for iron deficiency anemia in pre-post method. Lastly, IMPRINT study provided food supplements and compared them with the oral iron group.

Within the children’s datasets 7 studies contributed to a total of 18 datasets (sample - 13,435). First study IMPRINT(61), contributed to a total of 8 dataset delivered interventions as supplement or food vehicle whereas others have delivered interventions as supplement or through fortification. Studies have administered ferrous sulphate as interventions along with food supplements and some were Ayush trials.

## Discussion

The PRAYAS cohort is a compilation of datasets from India on Anemia amongst women and children. This compilation is in response to prolonged deliberations regarding stagnancy in the prevalence of anemia in India despite focused interventions like AMB. Studies have explained an increase in the compliance to such programmatic interventions that can accelerate reductions in anaemia prevalence (63). Despite such decisive interventions and framework, findings from Nationally representative sample surveys highlights an increase in anemia prevalence among WRA (from 53.1% in 2015–16 to 59.1% in 2019–21), pregnant women (from 50.3% to 52.6%), and children aged 6–59 months (from 58.6% to 67.1%) over the same period in India (11). Another study noted that there is an obvious shift in the distribution of Hb to the right among pregnant women over the past several years (28). This shift could be attributed to implementation of the programmatic interventions with a focus on pregnant women or to factors stemming from overall development. Dearth of robust evidence around the diverse clinical etiologies of anaemia, effective interventions etc. demands a study that can be used for further policy decision-making.

### Strengths and limitations

Data synthesized from the pooled data cohort would be used for calculating anemia indicators for the given population as these data have been collected from high quality and closely observed observational and randomized controlled studies mostly using venous blood samples. This is an important resource considering several challenges associated with the existing health surveys (12–14). Additionally, the analysis would provide etiological fractions for anemia prevalence importantly fraction due to iron deficiency in all age groups children under 18 years, NPNL, and pregnant women. These details can help the program to decide on the necessity of continuing prophylactic supplementation for these age groups and also finetune the doses for the same. Also, the individual patient data meta-analysis of intervention studies can inform robust evidence regarding the type of iron intervention, dose of iron in both therapeutic and prophylactic studies.

The analysis from this cohort profile is expected to generate robust, high-quality evidence from large high-quality studies to inform public health policies and guide strategies for reducing anemia’s burden in India. The systematic harmonization approach employed in this study ensures the validity and reliability of the datasets by addressing variations in data collection and standardizing outcome measures. This methodological rigor will enable more precise estimates and facilitate meaningful comparisons across populations and interventions (24–26).

However, several limitations should be noted. First, pooling data from studies with varying intervention types may result in high heterogeneity, which will be addressed through subgroup and sensitivity analyses. Second, some studies may lack critical parameters needed to assess changes in haemoglobin concentration, limiting the scope of certain analyses. Additionally, challenges in obtaining participant-level data due to restrictions from principal investigators or unpublished results could lead to data gaps. The inclusion of heterogeneous intervention and control conditions may also introduce a risk of bias, complicating the generalizability of findings. To mitigate these issues, we will evaluate heterogeneity using advanced statistical models, such as random-effects meta-analysis, and conduct subgroup analyses to explore the impact of differences across geographic, demographic, and intervention-specific factors.

The ability to analyse participant-level data allows for greater flexibility in adjusting for confounders, exploring effect modifiers, and conducting tailored subgroup analyses. By addressing sources of heterogeneity and potential biases, this meta-analysis aims to provide nuanced and reliable insights into the epidemiology of anemia and the effectiveness of various interventions.

Analysis from this cohort profile will contribute significantly to understanding the factors driving anemia’s high prevalence in India and the efficacy of interventions aimed at combating this public health challenge. The findings will support evidence-based policymaking i.e. will feed into the Anemia Mukt Bharat program recommendations for WRA and children and guide targeted strategies to reduce anemia and its associated health burdens across vulnerable populations.

## FURTHER DETAILS

### Study dissemination

Study findings will be published in peer-reviewed journals and will also be communicated with the policy makers for effective decision making to curb the increasing trend of anemia in India.

### Data availability statement

All the collaborating PIs have acknowledged that the pooled data can only be used for this IPD analysis, with no transfer of ownership.

## Acknowledgement

None

## Contributors

Conceptualization (C) & Study Design (SD) ; Data Collection (DC) & Curation (C) ; Data Analysis (DA) & Interpretation (I); Manuscript Writing (MW) & Revision (R) ; Supervision (S) & Project Administration (PA)

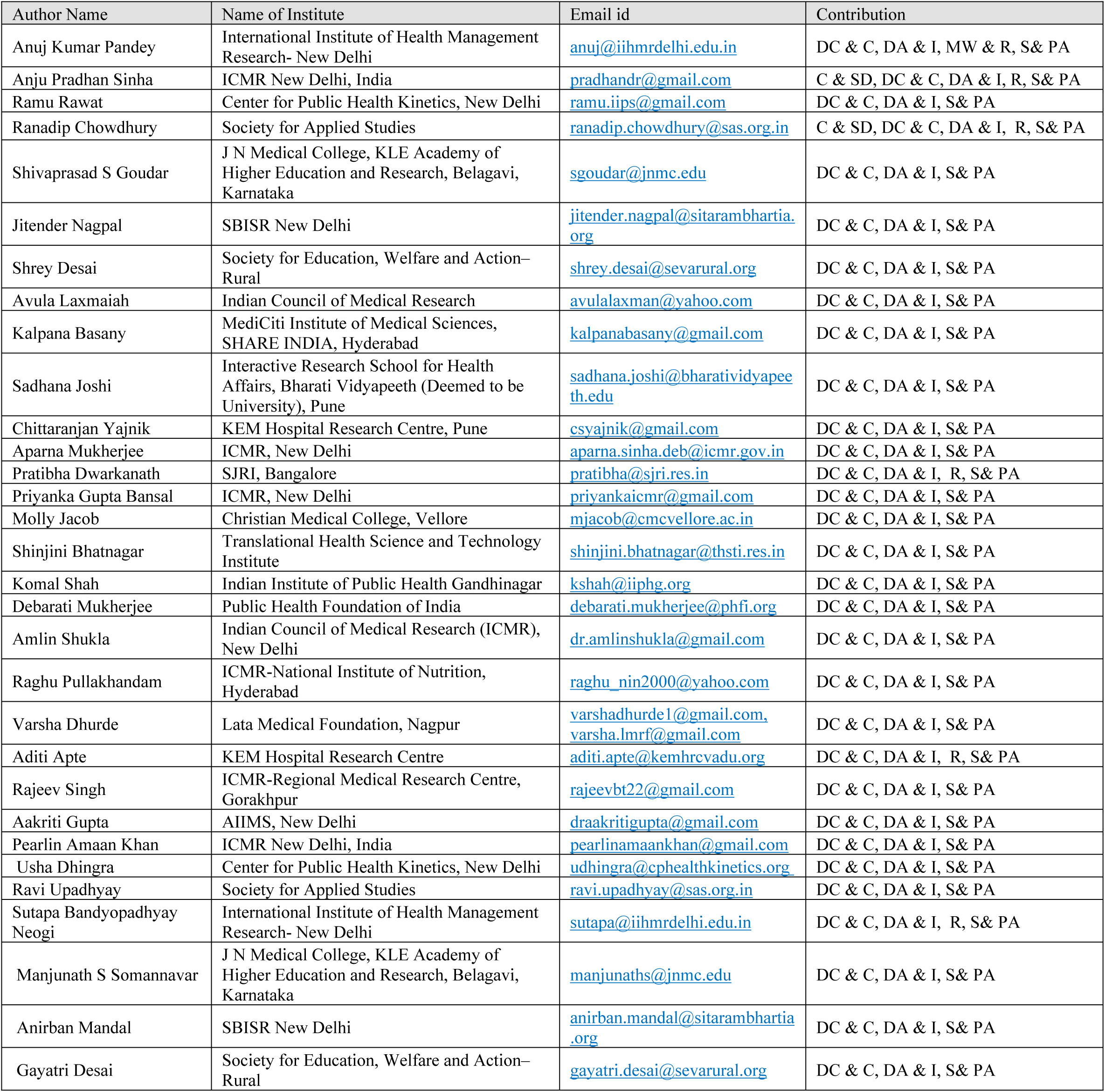
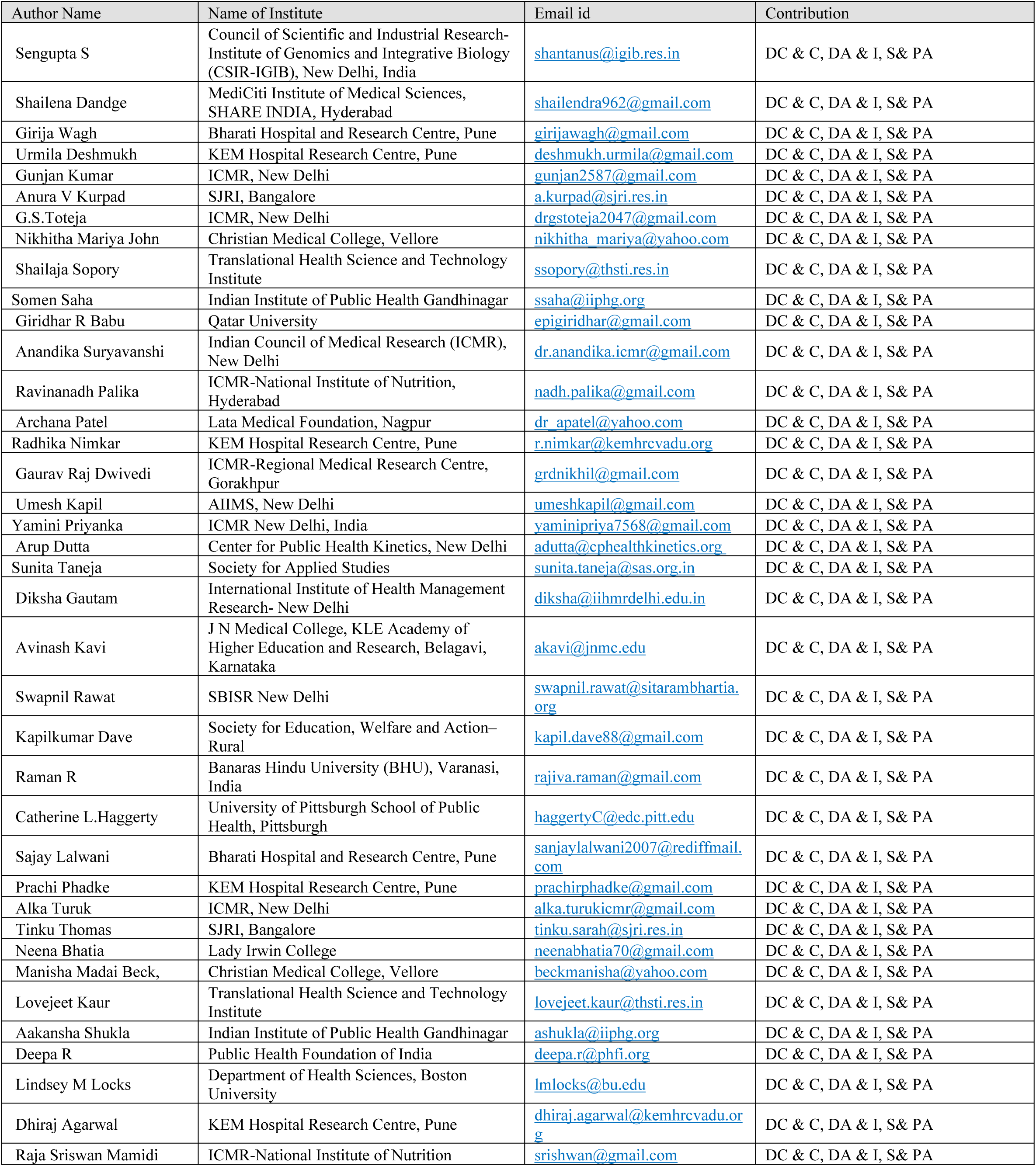
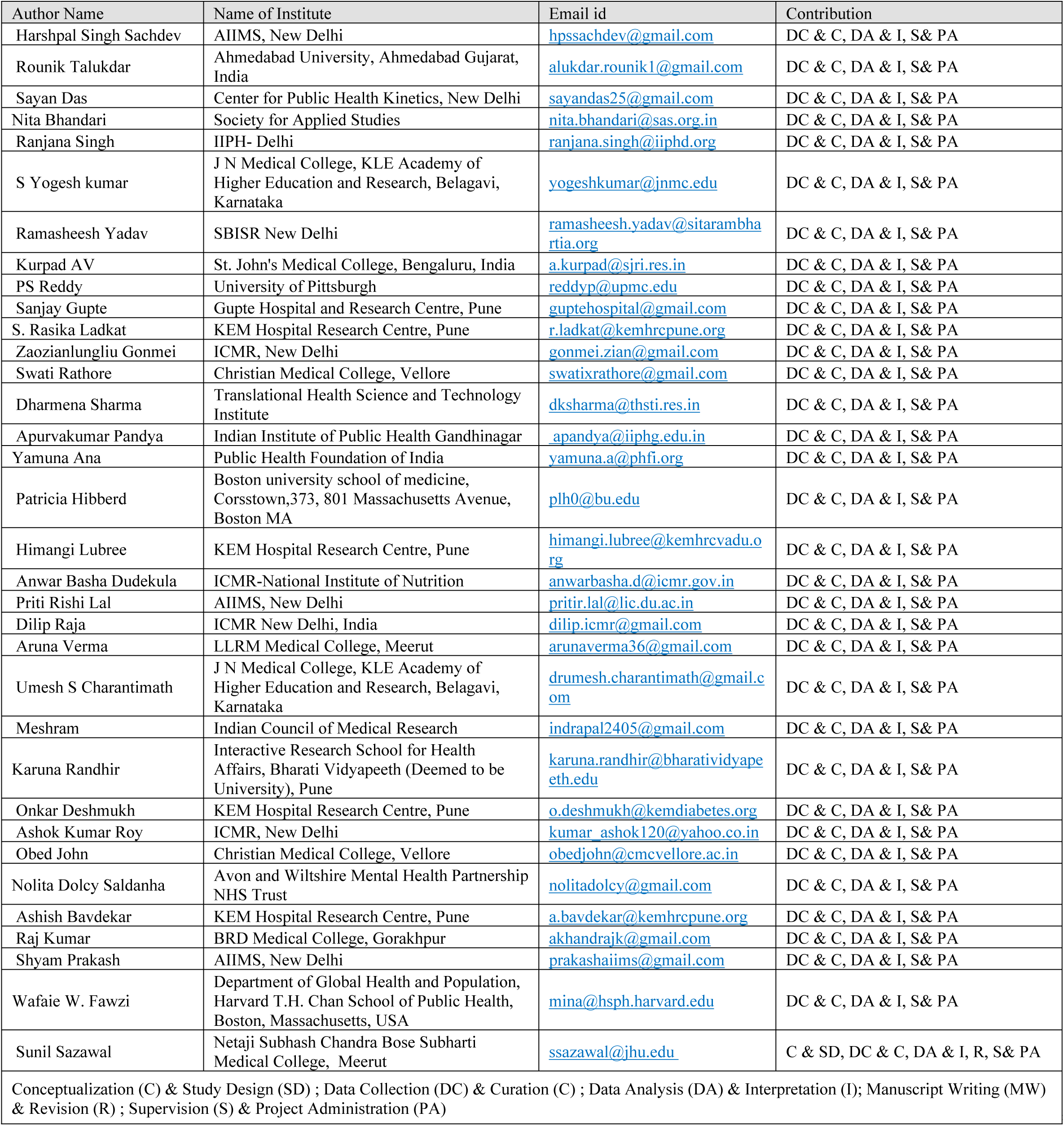

## Provenance and peer review

Commissioned by Indian Council of Medical Research – India

